# Does a probiotic (L. reuteri) lozenge taken twice daily over 3–4 weeks reduce probing pocket depth in patients with chronic periodontitis after 3 months? A systematic review of clinical trials (Preprint)

**DOI:** 10.1101/2023.05.15.23289010

**Authors:** Steffen Mickenautsch, Stefan Rupf, Veerasamy Yengopal

**Affiliations:** Review Centre for Health Science Research, 84 Concorde Road East, Bedfordview/Johannesburg, 2008, South Africa; Department of Community Dentistry, School of Oral Health Sciences, Faculty of Health Sciences, University of the Witwatersrand, 7 York Rd, Parktown/Johannesburg, 2193, South Africa; Synoptic Dentistry, Saarland University, Building 73, 66421, Homburg, Germany; Faculty of Dentistry, University of the Western Cape, Francie van Zijl Avenue, Tygerberg/Cape Town, 7505, South Africa

## Abstract

**Introduction:** Chronic periodontitis is a slow-progressing, multifactorial inflammatory disease of the periodontium that may lead to its destruction, which is detectable as increasing probing pocket depth (PPD), subsequent tooth mobility and tooth loss. The purpose of this systematic review was to update and appraise the current trial evidence to the question: Does probiotic (L. reuteri) lozenge taken twice daily over 3–4 weeks reduce PPD in patients with chronic periodontitis after 3 months?

**Methods and analysis:** Reference checks of previous systematic review and trial reports on the topic were conducted. PubMed, Scopus, Cochrane Library and the Directory of Open Access Journals (DOAJ) were searched. All selected trial reports were independently appraised by two reviewers, using the CQS-2B trial appraisal tool. Meta-analysis was conducted using a random effect model with the inverse variance method, stratified according to CQS-2B corroboration levels (C1–C4). The I^2^-test with 95% confidence interval was used to establish whether any statistical heterogeneity exists between datasets.

**Results:** Seven trials were included for appraisal and analysis. None complied with all appraisal criteria and thus were rated with an overall 0-score (high bias risk). Meta-analysis results at the lower C3-level (MD -0.64, 95% CI: -1.09 to -0.18) were found as being at risk of overestimating the true probiotic effect.

**Conclusion:** The clinical evidence identified in this systematic review is at high risk of representing an overestimation of the true therapeutic effect due to systematic error. The results of large randomised control trials are needed before any clinically relevant answer to the review question can be made.

## Introduction

Chronic periodontitis is a slow-progressing, multifactorial inflammatory disease of the periodontium that may lead to its destruction, which is detectable as increasing probing pocket depth (PPD), subsequent tooth mobility and tooth loss [1]. Treatment in its early stage comprises non-surgical periodontal therapy (NSPT), particularly scaling and root planning (SRP) [2].

A comprehensive systematic review by Ausenda et al. (2023) appraised the clinical evidence regarding the possible beneficial effect of the adjunctive use of a probiotic for chronic periodontitis treatment. The authors appraised the clinical evidence for the type of probiotic application (via lozenge, capsule or other); application frequency (1 or 2 times per day); type of probiotic (Lactobacillus reuteri, others), type of measured outcome PPD reduction, clinical attachment level (CAL) gain) and the length of the follow-up period (< 3, 3–12, > 12 months) [2]. The result of the systematic review showed that besides the application of NSPT, the intake of a probiotic lozenge, containing L. reuteri, twice daily for a period of 3–4 weeks in comparison to placebo was associated with the highest treatment benefit in terms of a statistically significant PPD reduction after a minimum period between 3–12 months. The point estimates of the mean differences (MD) with a 95% confidence interval (CI) for PPD reduction were higher and thus even more promising than that of the CAL gain for the same type of probiotic, application type and frequency [2]. An earlier second systematic review by Song and Liu (2020) reported similar results [3].

Both systematic reviews appraised clinical trials using the first version of Cochrane’s Risk of Bias (RoB) tool [4]. While most of the trials were judged to be of “low bias risk”, some were rated as moderate and high risk due to lack of adequate randomisation and blinding [2,3]. However, both systematic reviews based their review conclusions solely on the established trial data, without neither stratification by overall risk-of-bias judgment nor any other form of quantitatively integrating the established bias risk into their clinically relevant conclusions. In addition, the first RoB version has been found to have low interrater reliability: kappa 0.54 (95% CI: 0.29–0.79) [5], kappa 0.50 (95% CI: 0.36–0.63); and low agreement across reviewer pairs: kappa 0.37 (95% CI: 0.19–0.55) [6], which carries the high risk that the results, established in both systematic reviews, may be subjective and not reviewer independent. For that reason, potential high bias risk may have affected an overestimation of the systematic review results and subsequently its clinically relevant conclusions.

In contrast, it has been shown that the Composite Quality Score (CQS) for the appraisal of prospective, controlled clinical therapy trials has a high interrater reliability: Brennan-Prediger coefficient (BPC) of 0.95; 95% CI: 0.87–1.00, which compared favourably to that of the first RoB tool version, with most of the differences between the RoB and the CQS being statistically significant (p < 0.05) in favour of the CQS [7]. In addition, the latest CQS version (CQS-2B), besides its high interrater reliability [8], has been established on a rigorous evidence base [9, 10] and sound epistemic principles [11], and is recommended to be applied together with stratification by overall risk-of-bias judgment of the established effect estimates [12].

The purpose of this systematic review was to update and, by using the CQS-2B tool, to appraise the current trial evidence for the most promising type of probiotic application for the treatment of chronic periodontitis [2] and thus to answer the question: Does a probiotic (L. reuteri) lozenge taken twice daily over 3–4 weeks reduce PPD in patients with chronic periodontitis after 3 months?

## Materials and methods

The protocol of this systematic review has been made available online on medRxiv [13] and has been registered with the International Prospective Register of Systematic Reviews (PROSPERO) under number CRD42023421014 prior to its start.

### Participants, intervention, comparison, outcome and study design (PICOS)

#### Participants (P)

Adult patients suffering from chronic periodontitis.

#### Intervention (I)

NSPT with additional administration of a probiotic lozenge containing L. reuteri, taken two times per day for a 3- to 4-week (21- to 28-day) period.

#### Comparison (C)

NSPT with placebo, taken two times per day for a 3- to 4-week (21- to 28-day) period.

#### Outcome (O)

Changes in pocket probing depth (PPD) after a minimum period of three months (up to a maximum of 12 months) after the start of treatment. The PPD is defined as the difference between the gingival margin and the bottom of the periodontal pocket, measured with a periodontal probe in millimetres (mm). The measure of effect for the PPD changes was set as the MD with 95% CI.

#### Study design (S)

Prospective, controlled clinical therapy trials with parallel group design.

### Systematic literature search

Reference checks of the two previous systematic review reports [2, 3], as well as of identified trials, for suitable trial reports were conducted. The search period for the systematic review by Song and Liu (2020) was between 2009 and 2019 [3] and the search cut-off date for the systematic review by Ausenda et al. (2023) was 5 March 2020 [2]. Therefore, it was assumed that the systematic literature search by the two reviews has identified all relevant trial reports published prior to 5 March 2020. In addition to the reference check, the databases searched were PubMed, Scopus, Cochrane Library and the Directory of Open Access Journals (DOAJ) using the string of search terms: chronic periodontitis AND lactobacillus reuteri. The search in PubMed was limited to 5 March 2020 to 25 April 2023. The search in Scopus was between 2020 and April 2023 and the search in the Cochrane Library between March 2020 and April 2023.

One reviewer (SM) conducted the searches by screening citation titles and abstracts and retrieved the full-text articles. A second reviewer (SR) independently verified the retrieved trial reports for eligibility. Any disagreements were resolved via discussion and consensus.

### Trial selection criteria

Published trial reports in any publication languages that complied with all the following criteria were eligible for selection:

i. Prospective, controlled clinical therapy trial;
ii. Trial characteristics in line with specified PICOS;
iii. Trial report published in full; and
iv. Computable continuous data for test and control group reported, including the total number of subjects, mean PPD values with standard deviation (SD) or standard error (SE).

Trial reports that during the review process were found not to comply with all criteria were excluded.

### Data extraction from accepted trials

All trial reports that were deemed relevant during the systematic literature search were traced in full copy and the following information was extracted:

i. Full reference details;
ii. Basic trial characteristics:
  a. Number of patients enrolled at baseline per intervention group;
  b. Mean patient age with SD per intervention group;
  c. Patients are smokers (Yes/No);
  d. Patients are Type II diabetics (Yes/No);
  e. Adjunctive antibiotic therapy provided (Yes/No);
iii. Computable data per intervention group.

One reviewer (SM) extracted all information and entered it into an MS Excel spreadsheet. A second reviewer (SR) double-checked the extracted data and corrected possible entry errors. Any disagreements were resolved via discussion and consensus.

### Main data analysis

Meta-analysis was conducted using a random effect model with an inverse variance method, stratified according to the four CQS-2B corroboration levels (C1–C4).

The I^2^-test with 95% CI was used to establish whether any statistical heterogeneity between datasets exists. Thresholds for I^2^ point estimates (in %) were used to interpret the test results: 0–40% = might not be important; 30–60% = may represent moderate heterogeneity; 50–90% = may represent substantial heterogeneity; 75–100% = considerable heterogeneity [14].

### Sensitivity analysis

It was planned to conduct sensitivity analysis for meta-analysis results of trials that were rated with 1-score at all four CQS-2B appraisal criteria, by excluding trials in which:

a. Patients were smokers;
b. Patients were Type II diabetics; and
c. An adjunctive antibiotic therapy was provided.

The results of the sensitivity analysis were to be compared to that of the main analysis to ascertain whether smoking, Type II diabetes and antibiotic therapy had any possible confounding effect on the established main results.

### Assessment of bias risk

All selected trial reports were independently appraised by two reviewers (SM and SR) using the CQS-2B tool (Table 1). Any disagreements between reviewers were resolved by discussion and consensus.

**Table 1.** CQS-2B appraisal criteria.

|  |  |
| --- | --- |
| Criterion I | 'Randomisation' for allocation to treatment groups is in some form reported in the text |
| Criterion II | Any assurance that the patient allocation to treatment groups according to the random sequence was applied by an independent agent or agency, not otherwise involved in the trial, is in some form reported in the text |
| Criterion III | Double-blinding or the blinding of at least two out of the three groups: trial participants trial personnel and trial outcome assessors in some form reported in the text |
| Criterion IV | The sample size of any particular treatment group reported in the trial is not less than N = 100 |

Application of the CQS-2B comprises: (i) binary trial report rating per appraisal criterion (Scores: 0 = No/invalid/falsified, 1 = Yes/corroborated); (ii) multiplication of all criterion scores to an overall appraisal score, and (iii) identification of invalid/falsified trial reports based on a zero overall appraisal score.

During CQS-2B application, several corroboration (C) levels are recognised. C-levels indicate the number of consecutive criteria that a trial has complied with (e.g. level C2 indicates compliance with Criterion I and II; level C3 indicates compliance with Criteria I, II and III) [11]. A corroboration level for a particular trial is reached before one criterion is rated with a 0-score or when all criteria are rated with a 1-score; for example, corroboration level C2: Criterion I and II = 1-score, Criterion III = 0-score; corroboration level C4: All criteria = 1-score. After a criterion has been rated with a 0-score, the C-level of a trial remains the same, even if a following criterion is rated with a 1-score, for example corroboration level C2: Criterion I and II = 1-score, Criterion III = 0-score, Criterion IV = 1-score.

An overall 1-score appraisal result indicates that a trial is ‘corroborated’, which means that during the appraisal process, no evidence was established in support of the assumption that its reported results are compromised by high bias risk. This does not mean that such evidence may not be identified during future appraisals with additional appraisal criteria. For that reason, a corroborated trial is not assumed to be of ‘low bias risk’ status.

For all allocated 1-scores, the appropriate verbatim quotes were extracted from the trial report and entered into a verbatim table, including page number / column / paragraph number / line number of the trial report.

Further statistical assessment for the possible selection bias of meta-analysis results, including data of at least four trials at CQS-2B corroboration level 4 (i.e. trials which have been rated with a 1-score for all four appraisal criteria), was intended using the statistical test presented by Hicks et al. [15]. In addition, all trials judged at corroboration level C4 were to be further appraised for any other type of possible error related to their individual trial characteristics.

If the number of selected trials was at least ten, quantitative assessment of publication bias risk by use of Egger’s regression [16] was planned.

## Results

### Systematic literature search and data extraction

A total of 43 citations were identified through database searches and an additional seven citations through reference checking (Figure 1). Of these, 37 citations were deleted after abstract screening, due to lack of relevance in regard to the PICOS question. Therefore, a total of 13 citations were included for full trial report tracing and further review. Six trial reports were excluded for the following reasons: type of probiotic application did not include the use of a lozenge (three trial reports); no use of a placebo in the control groups (two trial reports) and one duplication of a trial report that was already included. Seven trial reports (reporting on six individual trials) were accepted for data extraction and trial appraisal [17–23]. All details of the systematic literature search results, including the references of the six excluded trial reports with reasons for exclusion, are presented in the Additional File / Section 1.

**Figure 1.**
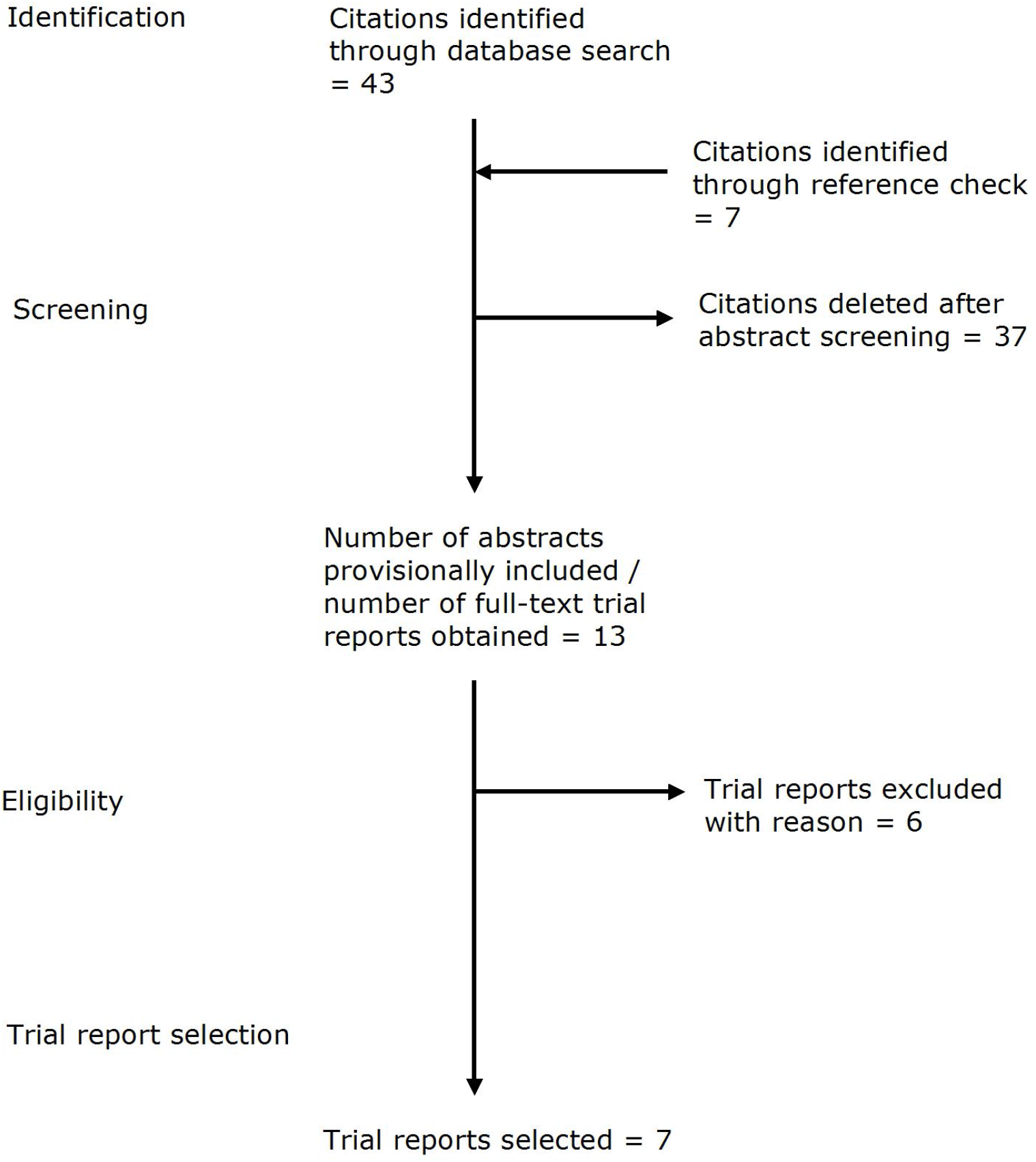
Flow diagram of the study selection process.

Two trial reports [17, 19] referred to the same registered trial (Ref no. UW16-043) but appeared to report treatment results of different patients in their test groups with the mean age of 52.30 (SD = 10.50) years [19] and 51.40 (SD = 8.88) years [17]. For that reason and in keeping with the previous systematic review by Ausenda et al. (2023) [2], both reports were included as separate trials.

The characteristics of the accepted trials are presented in Table 2. None of these included patients with Type II diabetes or antibiotic treatment. Only one trial [22] included smokers. The treatment duration in five trials [17, 19, 20, 22, 23] were 3, in one it was 12 [18] and in another one it was 16 weeks [21]. Patient follow-up varied between 3 and 13 months (Table 2).

**Table 2.** Characteristics of appraised trials.

| Trial | Patients |  |  | Test group |  |  | Control group |  |  | Treatment duration (in weeks) | Follow-up (in months) |
| --- | --- | --- | --- | --- | --- | --- | --- | --- | --- | --- | --- |
|  | Diabetes Type II | Smoker | Antibiotic treatment | n | Mean age | Age (SD) | n | Mean age | Age (SD) |  |  |
| Ince et al. 2015 | N | N | N | 15 | 41.00 | 3.17 | 15 | 42.20 | 2.78 | 3 | 13 |
| Laleman et al. 2019 | N | N | N | 20 | 58.00 | 12.00 | 19 | 58.00 | 13.00 | 12 | 6 |
| Pelekos et al. 2019 | N | N | N | 21 | 52.30 | 10.50 | 20 | NR | NR | 3 | 6 |
| Pelekos et al. 2020 | N | N | N | 20 | 51.40 | 8.88 | 20 | 52.76 | 6.67 | 3 | 6 |
| Tekce et al. 2015 | N | N | N | 20 | 43.00 | 5.01 | 20 | 41.40 | 8.8 | 3 | 13 |
| Theodoro et al. 2019 | N | Y | N | 14 | 47.25 | 7.10 | 14 | 45.07 | 6.31 | 3 | 3 |
| Mani et al. 2017 | N | N | N | 20 | 39.70 | 5.08 | 20 | 40.00 | 5.00 | 16 | 4 |

N/Y = No/Yes; n = Number of patients; SD = Standard deviation; NR = Not reported.

### Appraisal of bias risk and stratified meta-analysis

All trials were appraised using the CQS-2B tool (Table 1). The appraisal results are shown in Table 3 and the text-based justification for the allocation of 1-scores per appraisal criterion is presented in a verbatim table (see Additional File / Section 2). None of the seven included trials complied with all appraisal criteria and were therefore rated with an overall 0-score. Five trials [17–20, 23] received 1-scores for Criterion I–III (= corroboration level 3); one trial [22] complied with Criteria I and II (= corroboration level 2) and one trial [21] with Criteria I and III (= corroboration level 1). All trials had less than 100 patients included per intervention group and thus did not comply with Criterion IV.

**Table 3.** Trial appraisal results.

| Trial | CQS-2B Criterion |  |  |  | Total score | Corroboration level |
| --- | --- | --- | --- | --- | --- | --- |
|  | I | II | III | IV |  |  |
| Ince et al. 2015 | 1 | 1 | 1 | 0 | 0 | C3 |
| Laleman et al. 2019 | 1 | 1 | 1 | 0 | 0 | C3 |
| Pelekos et al. 2019 | 1 | 1 | 1 | 0 | 0 | C3 |
| Pelekos et al. 2020 | 1 | 1 | 1 | 0 | 0 | C3 |
| Tekce et al. 2015 | 1 | 1 | 1 | 0 | 0 | C3 |
| Theodoro et al. 2019 | 1 | 1 | 0 | 0 | 0 | C2 |
| Mani et al. 2017 | 1 | 0 | 1 | 0 | 0 | C1 |

Score 0 = Trial result falsified / Score 1 = corroborated

The results of the meta-analysis, stratified by corroboration level (including the number of included patients and the MDs with SD in PPD per intervention group per trial) are presented in Figure 2. The pooled effect estimate (MD -0.64, 95% CI: -1.09 to -0.18) of the highest reached Level C3 – and thus the result with the least bias risk – which indicates a statistically significantly more reduced PPD for NSPT with probiotic treatment than with placebo.

**Figure 2.**
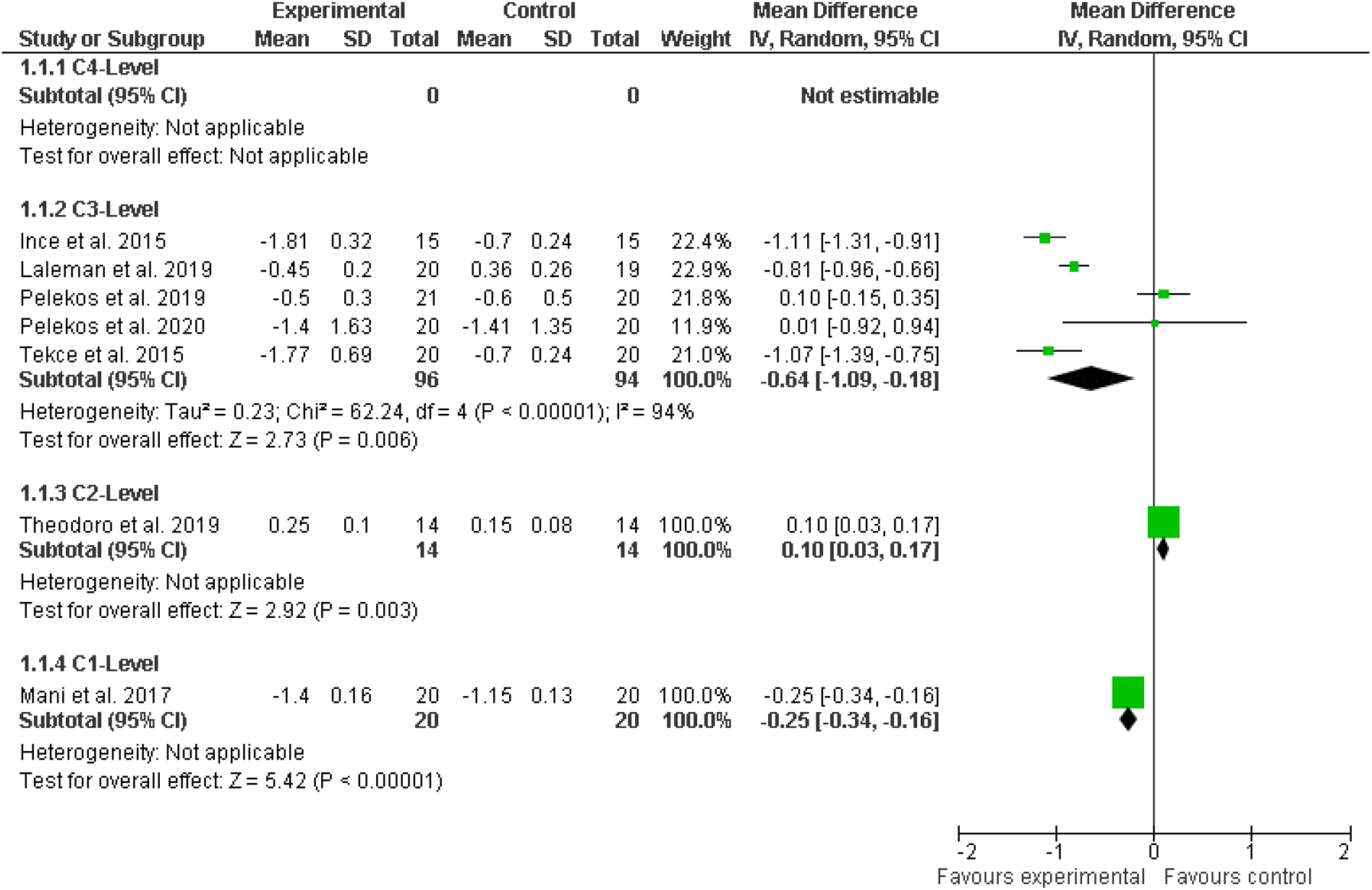
Meta-analysis results stratified by corroboration (C) levels (overall bias risk)

Based on the established bias risk appraisal result (Table 3), the pooled effect estimate at C3 Level (Figure 2) is at high risk of being an overestimation of the true therapeutic value. All included trials are small studies and thus may be affected by a 33% overestimation. Such a high risk of systematic error has been observed to be associated with therapy trials that have less than 100 patients included per intervention group [9]. When such overestimation is factored into the established meta-analysis result, the pooled effect estimate (MD -0.64, 95% CI: -1.09 to -0.18) changes into a statistically non-significant result (MD 0.14, 95% CI: -0.46 to 0.75).

In addition, a considerable statistical in-between trial heterogeneity (I^2^ = 94%) was observed for the pooled effect estimate at C3 Level (Figure 2). Since the meta-analysis was conducted by using a random-effects model, the possibility that the heterogeneity was simply due to chance was excluded. Therefore, possible reasons were explored by the step-wise exclusion of single trials from the meta-analysis. It was found that by excluding the two trials by Pelekos et al. [17, 19] and the trial by Laleman et al. (2019) [18], the heterogeneity (I^2^) of the pooled effect estimate of the two remaining trials (MD -1.10, 95% CI: -1.27 to -0.93) [20, 23] reached zero value. It was also noted that even when a 33% overestimation was factored into this result, the adjusted pooled effect estimate remained statistically significant in favour of probiotic treatment (MD -0.47, 95% CI: -0.70 to -0.25). Since none of the trials reached Level C4, no further bias risk assessment was conducted.

Because the included number of trials was < 10, no publication bias was assessed. We also did not undertake sensitivity analysis, because only one trial by Theodoro et al. (2019) [22] included smokers that were not pooled with any other trials at its reached corroboration level (Figure 2).

## Discussion

The aim of this systematic review was to answer the question of whether a probiotic (L. reuteri) lozenge taken twice daily over 3–4 weeks reduces PPD in patients with chronic periodontitis after 3 months.

The currently available clinical evidence does not provide a sufficient basis for answering this question. The pooled effect estimate of identified trials with the least risk of bias at Level C3 (MD - 0.64, 95% CI: -1.09 to -0.18, I^2^ = 94%) provides a good justification for the hypothesis that a probiotic (L. reuteri) lozenge may be effective as adjunct in the treatment of chronic periodontitis. However, the high risk of systematic error (bias), due to a too low sample size, indicates a high probability that this estimate represents a significant overestimation and that the true therapeutic effect might be not more beneficial than that of a placebo (MD 0.14, 95% CI: -0.46 to 0.75).

In addition, when the considerable in-between trial heterogeneity (I^2^ = 94%) of the pooled effect estimate was explored by excluding the two trials by Pelekos et al. [17, 19] and the trial by Laleman et al. (2019) [18], a robust pooled effect estimate from the two trials by Tekce et al. [20] and Ince et al. (2015) [23] was established (MD -1.10, 95% CI: -1.27 to -0.93, I^2^ = 0%). The reason may be that both trials presented results at a follow-up period of 13 months, while all other trials [17–19] presented results after a follow-up of 6 months only (Table 2). The difference between the non-significant effect estimates of the two trials by Pelekos et al. [17, 19] and the statistically significant effect estimate by Laleman et al. (2019) [18] may be explained by the fact that the treatment duration of the former was 3 months and that of the latter was 12 weeks long (Table 2).

These observations may provide support for the hypotheses that a three-week probiotic treatment may have a beneficial effect after 13 months [20, 23] and a 12-week treatment a beneficial effect after a six-month follow-up period above that of any placebo effect [18]. However, these hypotheses require rigorous testing through large randomised control trials (RCT) with a sample size > 100 patients per intervention group before any clinically relevant answer to the review question can be made.

The results of this systematic review confirm the assessment of the European Food Safety Authority (EFSA), which stated that the current evidence is insufficient to establish a cause-and-effect relationship between the consumption of a probiotic (L. reuteri) lozenge and the maintenance of normal gum function [24]. In contrast, the results are in disagreement with the systematic review findings by Ausenda et al. (2023) that the use of probiotics provides an additional benefit as an adjunct to NSPT in patients with periodontitis [2].

Six [17–21, 23] out of the seven clinical trials included in our review were identified during reference checking of the systematic review by Ausenda et al. (2023) and none was further identified during our database search. Therefore, the reason for the difference in systematic review results may be ascribed to a number of methodological differences. In our systematic review, we stratified our meta-analysis results by level of bias risk in accordance with published recommendations [25]. Furthermore, based on meta-epidemiological study evidence [9], we considered the potential impact that systematic error may have had on the meta-analysis results and we explored potential reasons for the measured considerable statistical in-between trial heterogeneity. In contrast, although Ausenda et al. (2023) appraised clinical trials for bias risk, neither stratification of the meta-analysis results nor any consideration of systematic error effect was included. While the authors did establish a considerable statistical in-between trial heterogeneity, particularly for PPD reduction due to L. reuteri probiotic after more than three months (I^2^ = 99%), no exploration of possible reasons for such heterogeneity was conducted [2].

A further difference between both systematic reviews was the choice of the clinical trial appraisal tool. Ausenda et al. (2023) [2] used the first version of Cochrane’s RoB tool [26], while we applied the latest version of the Composite Quality Score (CQS-2B) [9, 10]. Although both tools assess randomisation and blinding in clinical trials, the CQS-2B also appraises trial sample size as a source for potential systematic error, which was not included in the RoB tool. Sample size limitations are commonly regarded as a problem concerning random error. Random error has been defined as the difference between the observed and the true value by chance, due to natural variations or imprecise measurement [27]. Such error affects trial precision, namely the reproducibility of results under the same conditions, with the probability of Type II – error (β) being of importance [9, 27]. This probability may be reduced on the basis of power calculation or when several small trials are combined by the use of meta-analysis. In contrast, a systematic review of meta-epidemiological studies established that clinical trials with a small sample size were also associated with effect overestimation due to systematic error [9]. Systematic error is the consistent deviation of the observed from the true value. It has been established that the inclusion of small trials may also divert the results of meta-analyses away from the true treatment effect size [28]. Because such error affects the accuracy of clinical trial results, namely how closely the observed trial results resemble the true ones – since this error appears to be consistent across a wide range of therapeutic interventions [9] – it was included as a criterion into the CQS-2B trial appraisal tool.

In summary, while this systematic review, similar to that by Ausenda et al. (2023) [2] and Song and Liu (2020) [3], also identified clinical trial data suggesting at first glance clinical benefit of the adjunct probiotic use for chronic periodontitis treatment, it could with the help of the CQS-2B tool integrate important trial characteristics concerning the risk of systematic error (bias) effect on such data into its review findings.

This further suggests that the use of the CQS-2B in systematic reviews of clinical therapy trials may, due to its high interrater reliability [8], rigorous evidence base [9, 10] and sound epistemic principles [11] applied in connection with recommended data stratification by overall risk-of-bias judgment [12], achieve generally more realistic appraisal results. The ability of this tool to identify elements that can potentially be a source of significant bias is important. With the CQS-2B, readers and authors of clinical trial reports will have an objective tool that they can use to determine the strength of the evidence used to arrive at a particular conclusion.

## Conclusion

The clinical evidence identified in this systematic review is at high risk of representing an overestimation of the true therapeutic effect. Although some of the clinical trial results provide support for the hypothesis that a 3-week probiotic treatment may have a beneficial effect after 13 months, and a 12-week treatment a beneficial effect after a 6-month follow-up period above that of any placebo effect, these hypotheses require rigorous testing through large RCT with a sample size > 100 patients per intervention group before any clinically relevant answer towards the review question can be made.

## Supporting information

PRISMA Checklist

PRISMA Flow Diagram

## Data Availability

All data produced in the present work are contained in the manuscript

## Data availability

All data have been made fully available without restriction as part of the preprint and journal publication.

